# Uncertainty Quantification in Radiogenomics: EGFR Amplification in Glioblastoma

**DOI:** 10.1101/2020.05.22.20110288

**Authors:** Leland S. Hu, Lujia Wang, Andrea Hawkins-Daarud, Jennifer M. Eschbacher, Kyle W. Singleton, Pamela R. Jackson, Kamala Clark-Swanson, Christopher P. Sereduk, Sen Peng, Panwen Wang, Junwen Wang, Leslie C. Baxter, Kris A. Smith, Gina L. Mazza, Ashley M. Stokes, Bernard R. Bendok, Richard S. Zimmerman, Chandan Krishna, Alyx B. Porter, Maciej M. Mrugala, Joseph M. Hoxworth, Teresa Wu, Nhan L. Tran, Kristin R. Swanson, Jing Li

## Abstract

**BACKGROUND:** Radiogenomics uses machine-learning (ML) to directly connect the morphologic and physiological appearance of tumors on clinical imaging with underlying genomic features. Despite extensive growth in the area of radiogenomics across many cancers, and its potential role in advancing clinical decision making, no published studies have directly addressed uncertainty in these model predictions.

**METHODS:** We developed a radiogenomics ML model to quantify uncertainty using transductive Gaussian Processes (GP) and a unique dataset of 95 image-localized biopsies with spatially matched MRI from 25 untreated Glioblastoma (GBM) patients. The model generated predictions for regional EGFR amplification status (a common and important target in GBM) to resolve the intratumoral genetic heterogeneity across each individual tumor - a key factor for future personalized therapeutic paradigms. The model used probability distributions for each sample prediction to quantify uncertainty, and used transductive learning to reduce the overall uncertainty. We compared predictive accuracy and uncertainty of the transductive learning GP model against a standard GP model using leave-one-patient-out cross validation.

**RESULTS:** Predictive uncertainty informed the likelihood of achieving an accurate sample prediction. When stratifying predictions based on uncertainty, we observed substantially higher performance in the group cohort (75% accuracy, n=95) and amongst sample predictions with the lowest uncertainty (83% accuracy, n=72) compared to predictions with higher uncertainty (48% accuracy, n=23), due largely to data interpolation (rather than extrapolation).

**CONCLUSION:** We present a novel approach to quantify radiogenomics uncertainty to enhance model performance and clinical interpretability. This should help integrate more reliable radiogenomics models for improved medical decision-making.

## INTRODUCTION

The field of machine-learning (ML) has exploded in recent years, thanks to advances in computing power and the increasing availability of digital data. Some of the most exciting developments in ML have centered on computer vision and image recognition, with broad applications ranging from e-commerce to self-driving cars. But these advances have also naturally dovetailed with applications in healthcare, and in particular, the study of medical images.^1^ The ability for computer algorithms to discriminate subtle imaging patterns has led to myriad ML tools aimed at improving diagnostic accuracy and clinical efficiency.^2^ But arguably, the most transformative application relates to the development of radiogenomics modeling, and its integration with the evolving paradigm of individualized oncology.^1–3^

Radiogenomics integrates non-invasive imaging (typically Magnetic Resonance Imaging, or MRI) with genetic profiles as data inputs to train ML algorithms. These algorithms identify the correlations within the training data to predict genetic aberrations on new unseen cases, using the MRI images alone. In the context of individualized oncology, radiogenomics non-invasively diagnoses the unique genetic drug sensitivities for each patient’s tumor, which can inform personalized targeted treatment decisions that potentially improve clinical outcome. Foundational work in radiogenomics coincided with the first Cancer Genome Atlas (TCGA) initiative over a decade ago,^4^ which focused on patients with Glioblastoma (GBM) - the most common and aggressive primary brain tumor. Since then, radiogenomics has extended to all major tumor types throughout the body,^5–8^ underscoring the broad scope and potential impact on cancer care in general.

But as clinicians begin to assimilate radiogenomics predictions into clinical decision-making, it will become increasingly important to consider the uncertainty associated with each of those predictions. In the context of radiogenomics, predictive uncertainty stems largely from sparsity of training data, which is true of all clinical models that rely on typically limited sources of patient data. This sparsity becomes an even greater challenge when evaluating heterogeneous tumors like GBM, which necessitate image-localized biopsies and spatially matched MRI measurements to resolve the regional genetic subpopulations that comprise each tumor. And as with any data driven approach, the scope of training data establishes the upper and lower bounds of the model domain, which guides the predictions for all new unseen test cases (i.e., new prospective patients). In the ideal scenario, the new test data will fall within the distribution of the training domain, which allows for interpolation of model predictions, and the lowest degree of predictive uncertainty. If the test data fall outside of the training domain, then the model must extrapolate predictions, at the cost of greater model uncertainty. Unfortunately, without knowledge of this uncertainty, it is difficult to ascertain the degree to which the new prospective patient case falls within the training scope of the model. This could limit user confidence in potentially valuable model outputs.

While predictive accuracy remains the most important measure of model performance, many non-medical ML-based applications (e.g.,weather forecasting, hydrologic forecasting) also estimate predictive uncertainty - usually through probabilistic approaches - to enhance the credibility of model outputs and to facilitate subsequent decision-making.^9^ In this respect, research in radiogenomics has lagged. Past studies have focused on accuracy (e.g., sensitivity/specificity) and covariance (e.g., standard error, 95% confidence intervals) in group analyses, but have not yet addressed the uncertainty for individual predictions.^10^ Again, such individual predictions (e.g. a new prospective patient) represent the “real world” scenarios for applying previously trained radiogenomics models. Quantifying these uncertainties will help to understand the conditions of optimal model performance, which will in turn help define how to effectively integrate radiogenomics models into effective clinical practice.

To address this critical gap, we present a probabilistic method to quantify the uncertainty in radiogenomics modeling, based on Gaussian Process (GP) and transductive learning.^11,9,12,13^ The GP method can quantify predictive uncertainty through probability distributions of regional genetic aberrations in each patient’s tumor. As a case study, we develop our model in the setting of GBM, which presents particular challenges for individualized oncology due to its profound intratumoral heterogeneity and likelihood for tissue sampling errors. We address the confounds of intratumoral heterogeneity by utilizing a unique and carefully annotated dataset of image-localized biopsies with spatially matched MRI measurements from a cohort of untreated GBM patients. As proof of concept, we focus on predictions of EGFR amplification status, as this serves as a common therapeutic target for many clinically available drugs. We demonstrate a progression of optimization steps that not only quantify, but also minimize predictive uncertainty, and we investigate how this relates to confidence and accuracy of model predictions. Our overarching goal is to provide a pathway to clinically integrating reliable radiogenomics predictions as part of decision support within the paradigm of individualized oncology.

## RESULTS

### Radiogenomics can resolve the regional heterogeneity of EGFR amplification status in GBM

We collected a dataset of 95 image-localized biopsies from a cohort of 25 primary GBM patients. We quantified and compared the EGFR amplification status for each biopsy sample with spatially matched image features from corresponding multi-parametric MRI, which included conventional and advanced (diffusion, tensor, perfusion) MRI techniques. We used these spatially matched datasets to construct a transductive learning GP model to classify EGFR amplification status. This model quantified predictive uncertainty for each sample, by measuring predictive variance and the distribution mean for each predicted sample, relative to the training model domain. We used these features to test the hypothesis that the sample belongs to the class predicted by the distribution mean (H1) versus not (H0), based on a standard one-sided z test. This generated a p-value for each sample, which reflected the uncertainty of the prediction, such that smaller p-values corresponded with lower predictive uncertainty (i.e., higher certainty). We integrated these uncertainty estimates using transductive learning to optimize model training and predictive performance. In leave-one-patient-out cross validation, the model achieved an overall accuracy of 75% (77% sensitivity, 74% specificity) across the entire pooled cohort (n=95), without stratifying based on the certainty of the predictions. **Figure 1** illustrates how the spatially resolved predictive maps correspond with stereotactic biopsies from the regionally distinct genetic subpopulations that can co-exist within a single patient’s GBM tumor.

**Figure 1:**
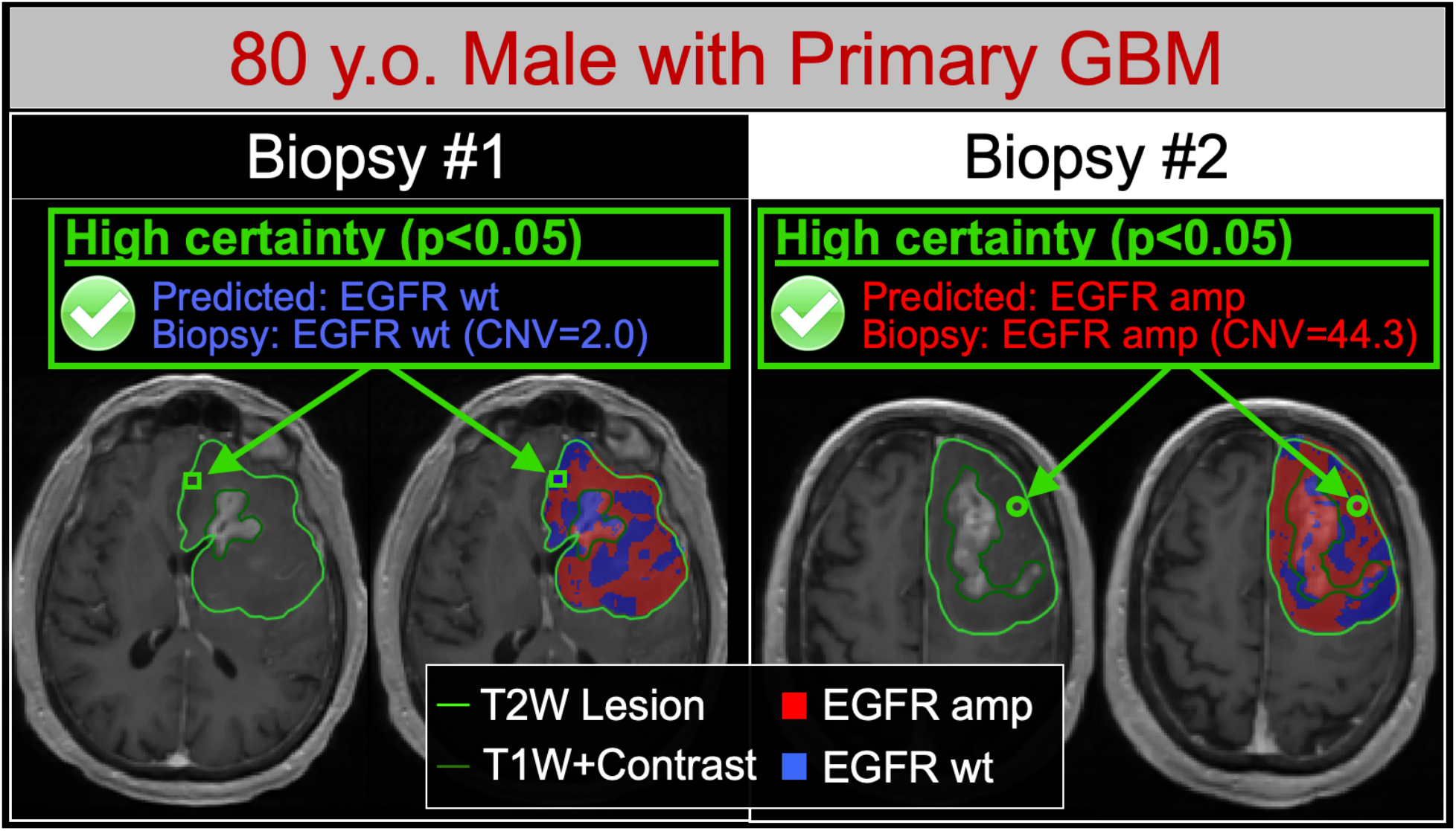
Radiogenomics maps resolve the regional intratumoral heterogeneity of EGFR amplification status in GBM. Shown are two different image-localized biopsies (Biopsy #1, Biopsy #2) from the same GBM tumor in a single patient. For each biopsy, T1+C images (left) demonstrate the enhancing tumor segment (dark green outline, T1W+Contrast) and the peripheral non-enhancing tumor segment (light green outline, T2W lesion). Radiogenomics color maps for each biopsy (right) also show regions of predicted EGFR amplification (amp, red) and EGFR wildtype (wt, blue) status overlaid on the T1+C images. For biopsy #1 (green square), the radiogenomics map correctly predicted low EGFR copy number variant (CNV) and wildtype status with high predictive certainty (p<0.05). Conversely for biopsy #2 (green circle), the maps correctly predicted high EGFR CNV and amplification status, also with high predictive certainty (p<0.05). Note that both biopsies originated from the non-enhancing tumor segment, suggesting the feasibility for quantifying EGFR drug target status for residual subpopulations that are typically left unresected followed gross total resection.

### Interpolation corresponds with lower predictive uncertainty and higher model performance

For data-driven approaches like ML, predictions can be either interpolated between known observations (i.e., training data) within the model domain, or extrapolated by extending model trends beyond the known observations. Generally speaking, extrapolation carries greater risk of uncertainty, while interpolation is considered more reliable. Our data suggest that prioritizing interpolation (over extrapolation) during model training can reduce the uncertainty of radiogenomics predictions while improving overall model performance.

As a baseline representation of current methodology, we trained a standard GP model without the transductive learning capability to prioritize maximal predictive accuracy for distinguishing EGFR amplification status, irrespective of the type of prediction (e.g., interpolation vs extrapolation). We applied this standard model to quantify which sample predictions in our cohort (n=95) were interpolated (p<0.05) versus extrapolated (p>0.05, standard deviation >0.4). When comparing with the standard GP model as the representative baseline, the transductive learning GP model shifted the type of prediction from extrapolation to interpolation in 11.6% (11/95) of biopsy cases. Amongst these sample predictions, classification accuracy increased from 63.6% with extrapolation (7/11 correctly classified) to 100% with interpolation (11/11 correctly classified).

The transductive learning GP model also reduced the number of sample predictions that suffered from uncertainty due to inherent imperfections of model classification. This specifically applied to those predictions with distributions that fell in close proximity to the classification threshold (so called border uncertainty), which associated with large p values (p>0.05) and low standard deviation (<0.40) (**Figure 2**). When comparing with the standard GP model as the representative baseline, the transductive learning GP model shifted the type of prediction, from border uncertainty to interpolation, in 15.8% (15/95) of biopsy cases. Amongst these sample predictions, classification accuracy dramatically increased from 40% in the setting of border uncertainty (6/15 correctly classified) to 93.3% with interpolation (14/15 correctly classified). **Figure 3** shows the overall shift in sample predictions when comparing the standard GP and transductive learning GP models, including the increase in interpolated predictions.

**Figure 2:**
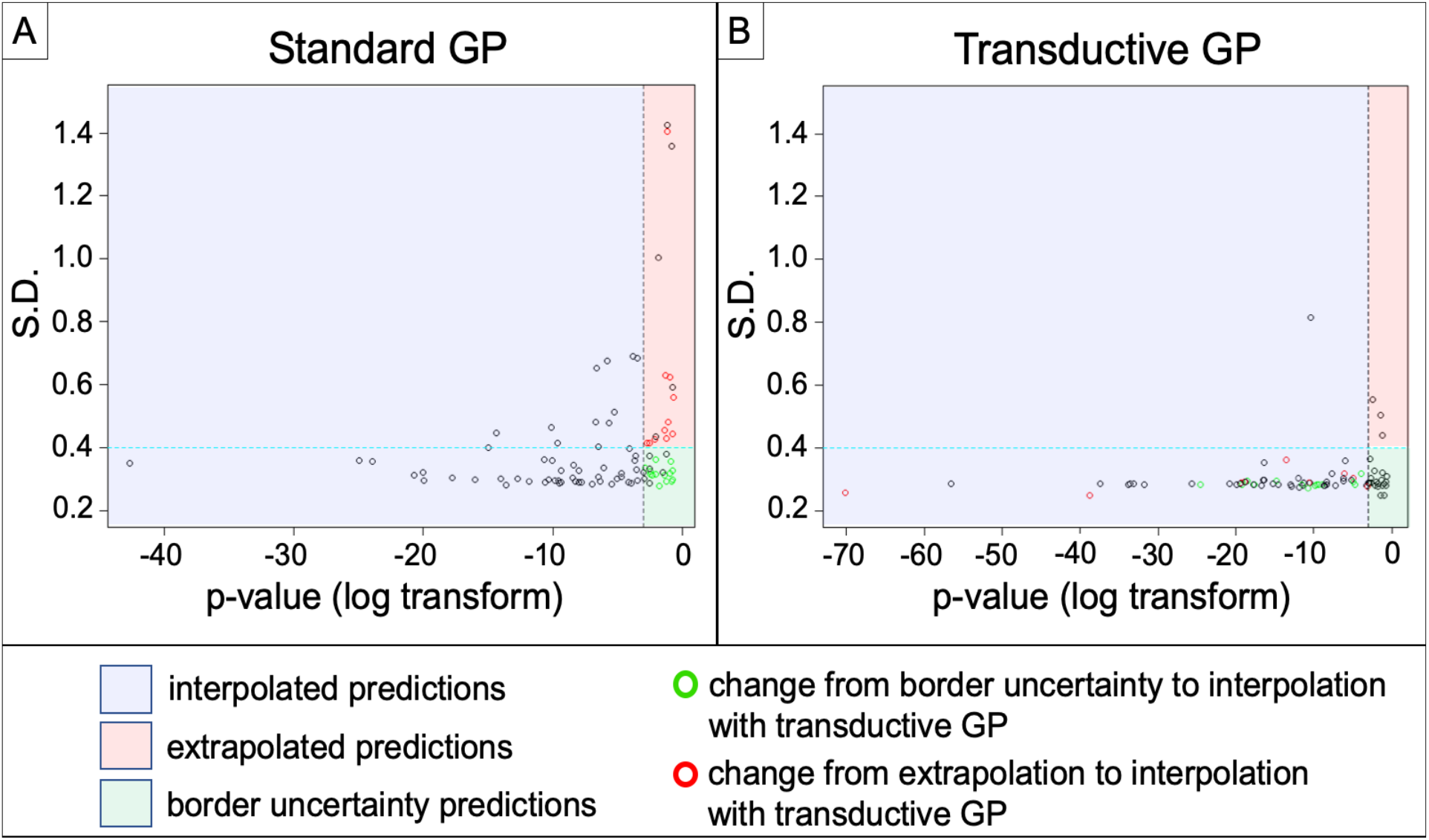
Transductive Learning GP increases the number of interpolated sample predictions compared to standard GP. Shown are scatter plots of standard deviation (S.D.) (y-axis) versus log-transform of p-value (x-axis) for all 95 sample predictions by the (A) Standard GP and (B) Transductive Learning GP models. The blue region demarcates those samples with p<0.05 (log transform < -3) corresponding to interpolated predictions. The red region demarcates extrapolated predictions (p>0.05, S.D. >0.40), while the green region demarcates predictions with border uncertainty (p>0.05, S.D. <0.40). The green circles denote those samples that shifted from border uncertainty to interpolated predictions with transductive learning. Similarly, the red circles denote those samples that shifted from extrapolated to interpolated predictions with transductive learning. Transductive GP produced 72/95 interpolated sample predictions, compared with 58/95 for Standard GP. Note the substantial decrease in extrapolated predictions with the transductive learning GP model.

**Figure 3:**
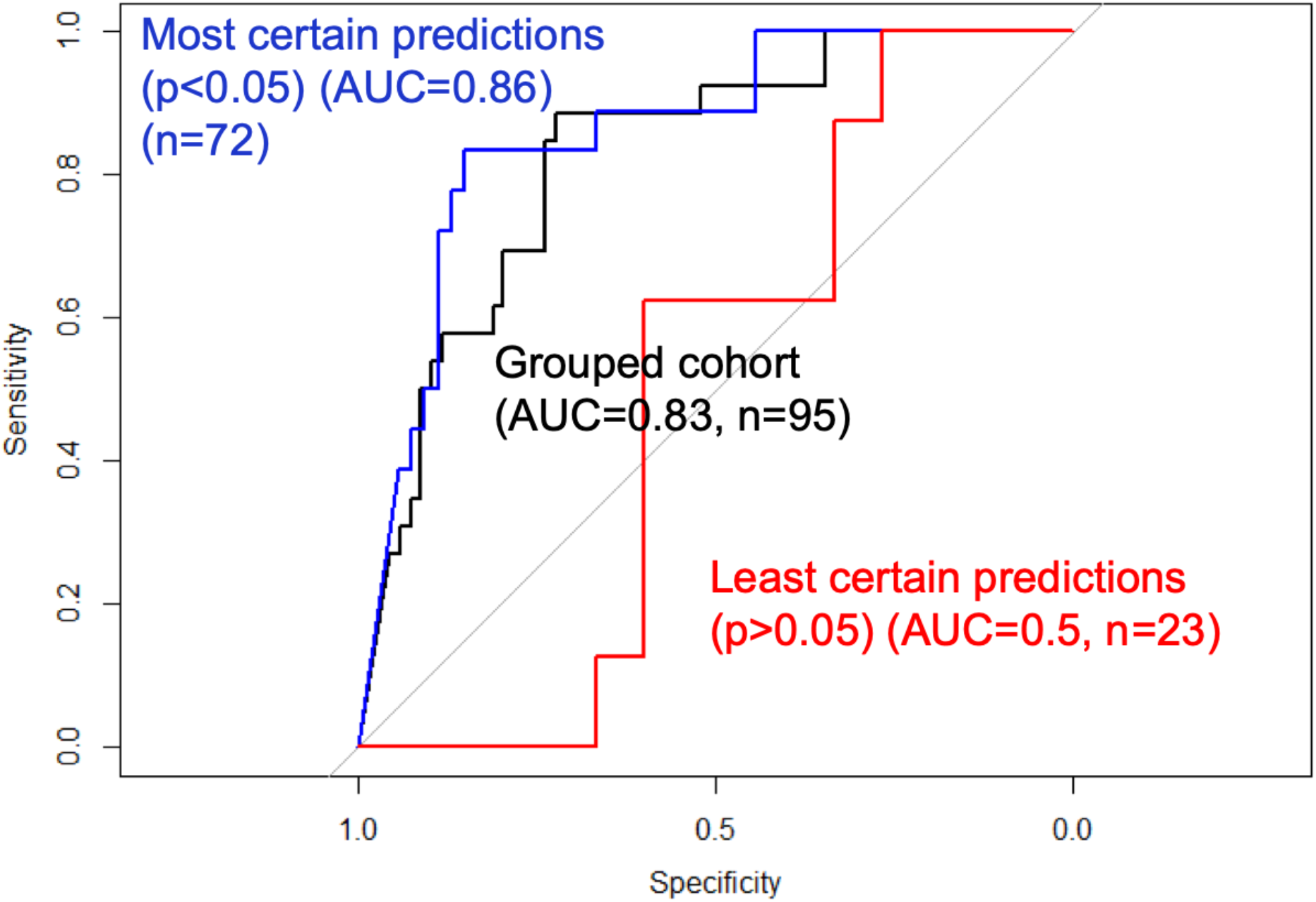
Model performance increases with lower predictive uncertainty. Shown are the Area-under-the-curve (AUC) measures for Receiver Operator Characteristics (ROC) analysis of sample predictions stratified by predictive uncertainty. The sample predictions with lowest uncertainty (p<0.05) (blue curve, n=72) achieved the highest performance (AUC=0.86) compared to the entire grouped cohort irrespective of uncertainty (AUC=0.83) (black curve, n=95). Meanwhile, the sample predictions with greatest uncertainty (i.e., least certain predictions) showed the lowest classification accuracy (AUC=0.5) (red curve, n=23).

We observed substantially different sizes in feature sets and overall model complexity when comparing the standard GP and transductive GP models, as summarized in **Table 1**. While the standard GP model selected 17 image features (across 5 different MRI contrasts), the transductive GP model selected only 4 features (across 2 MRI contrasts). The lower complexity of the transductive GP model likely stemmed from a key difference in model training: the transductive learning GP model first prioritized feature selection that minimized average predictive uncertainty (i.e., lowest sum of p-values), which helped to narrow candidate features to a relevant and focussed subset. Only then did the model prioritize predictive accuracy, within this smaller feature subset. Meanwhile, the standard GP model selected from the entire original feature set to maximize predictive accuracy, without constraints on predictive uncertainty. Although the accuracy was optimized on training data, the standard GP model could not achieve the same level of cross validated model performance (60% accuracy, 31% sensitivity, 71% specificity) compared to the transductive learning GP model, due largely to lack of control of extrapolation risks.

**TABLE 1:** Differences in Image features and model complexity when comparing standard GP and transductive learning GP models. The transductive learning model, which prioritized model uncertainty during the training process, comprised fewer image features and lower complexity compared with the standard GP model.

|  | Transductive Learning GP model | Standard GP model |
| --- | --- | --- |
| **Select ed image texture features** | 1. T2.Information.Measure.of.Correlation.2_Avg_1<br>2. T2.Angular.Second.Moment_Avg_1<br>3. T2.Kurtosis<br>4. rCBV.Contrast_Avg_1 | 1. T2.Difference.Entropy_Avg_3<br>2. T2.Contrast_Avg_1<br>3. T2.Entropy_Avg_1<br>4. SPGRC.Sum.Variance_Avg_3<br>5. SPGRC.Gabor_Std_0.4_0.1<br>6. rCBV.Angular.Second.Moment_Avg_1<br>7. rCBV.Difference.Variance_Avg_1<br>8. rCBV.Kurtosis<br>9. EPI.Gabor_Mean_0.4_0.1<br>10. EPI.Angular.Second.Moment_Avg_1<br>11. FA.Angular.Second.Moment_Avg_1<br>12. FA.Skewness<br>13. FA.Difference.Variance_Avg_3<br>14. FA.Entropy_Avg_1<br>15. FA.Sum.Variance_Avg_1<br>16. FA.Information.Measure.of.Correlation.1_Avg_1<br>17. FA.Range" |

### Predictive uncertainty can inform the likelihood of achieving an accurate sample prediction, which enhances clinical interpretability

Existing published studies have used predictive accuracy to report model performance, but have not yet addressed model uncertainty. Our data suggest that leveraging both accuracy and uncertainty can further optimize model performance and applicability. When stratifying transductive learning GP sample predictions based on predictive uncertainty, we observed a striking difference in model performance. The subgroup of sample predictions with the lowest uncertainty (i.e., the most certain predictions) (p<0.05) (n=72) achieved the highest predictive performance (83% accuracy, 83% sensitivity, 83% specificity) compared to the entire cohort as a whole (75% accuracy, n=95). This could be explained by the substantially lower performance (48% accuracy, 63% sensitivity, 40% specificity) observed amongst the subgroup of highly uncertain sample predictions (p>0.05) (n=23). **Figure 3** shows the differences in model performance on ROC analysis when comparing sample predictions based on uncertainty. These discrepancies in model performance persisted even when stratifying with less stringent uncertainty thresholds (e.g., p<0.10, p<0.15), which we summarize in **Table 2**. Together, these results suggest that predictive uncertainty can inform the likelihood of achieving an accurate sample prediction, which can help discriminate radiogenomics outputs, not only across patients, but at the regional level within the same patient tumor (**Figure 4**).

**TABLE 2:**
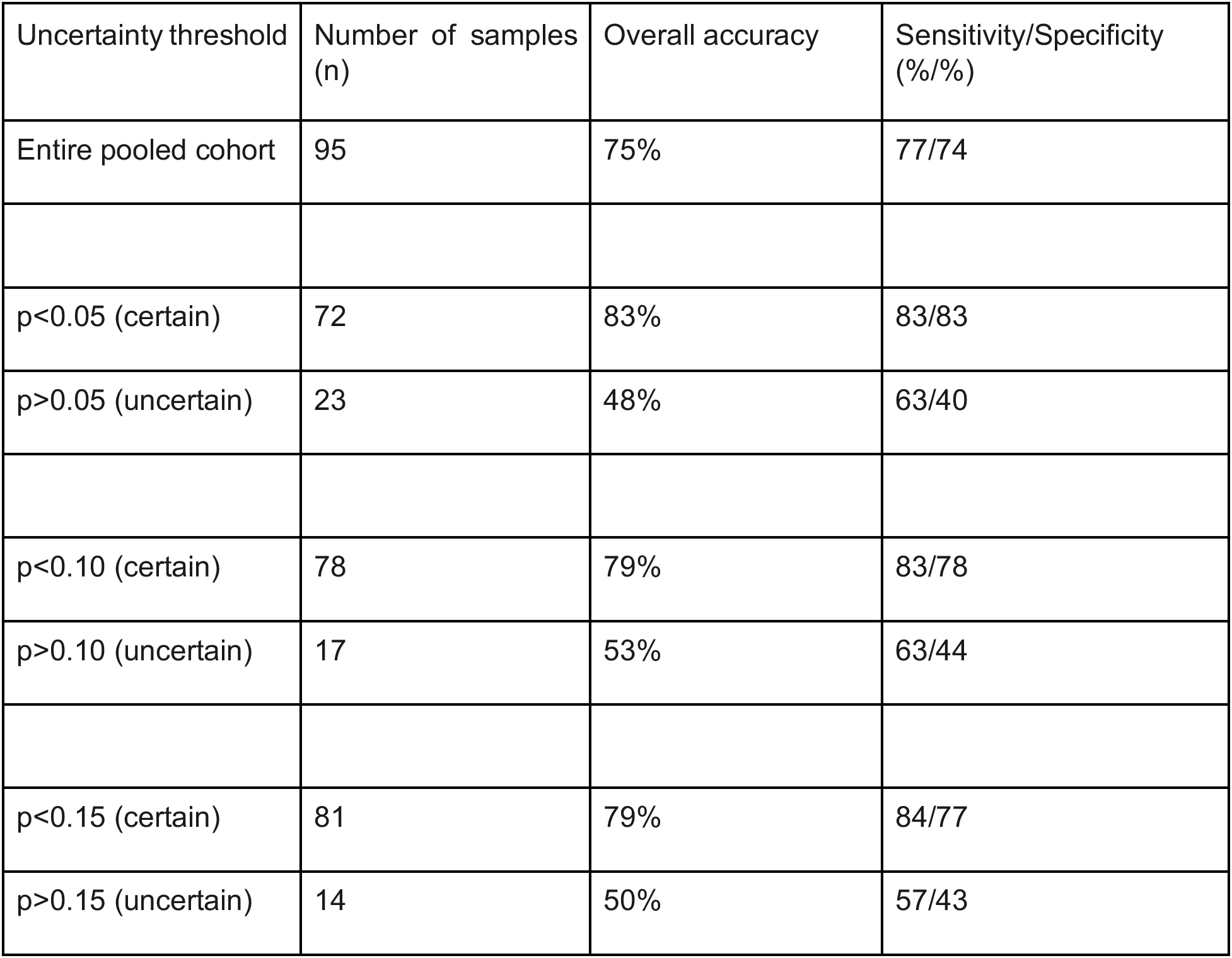
Differences in predictive accuracy related to certain versus uncertain sample predictions. Shown are the differences in predictive performance of the transductive learning GP model for the entire pooled cohort and also when stratifying the sample predictions with high certainty (low p-values) versus low certainty (high p-values) at various p-value thresholds.

**Figure 4:**
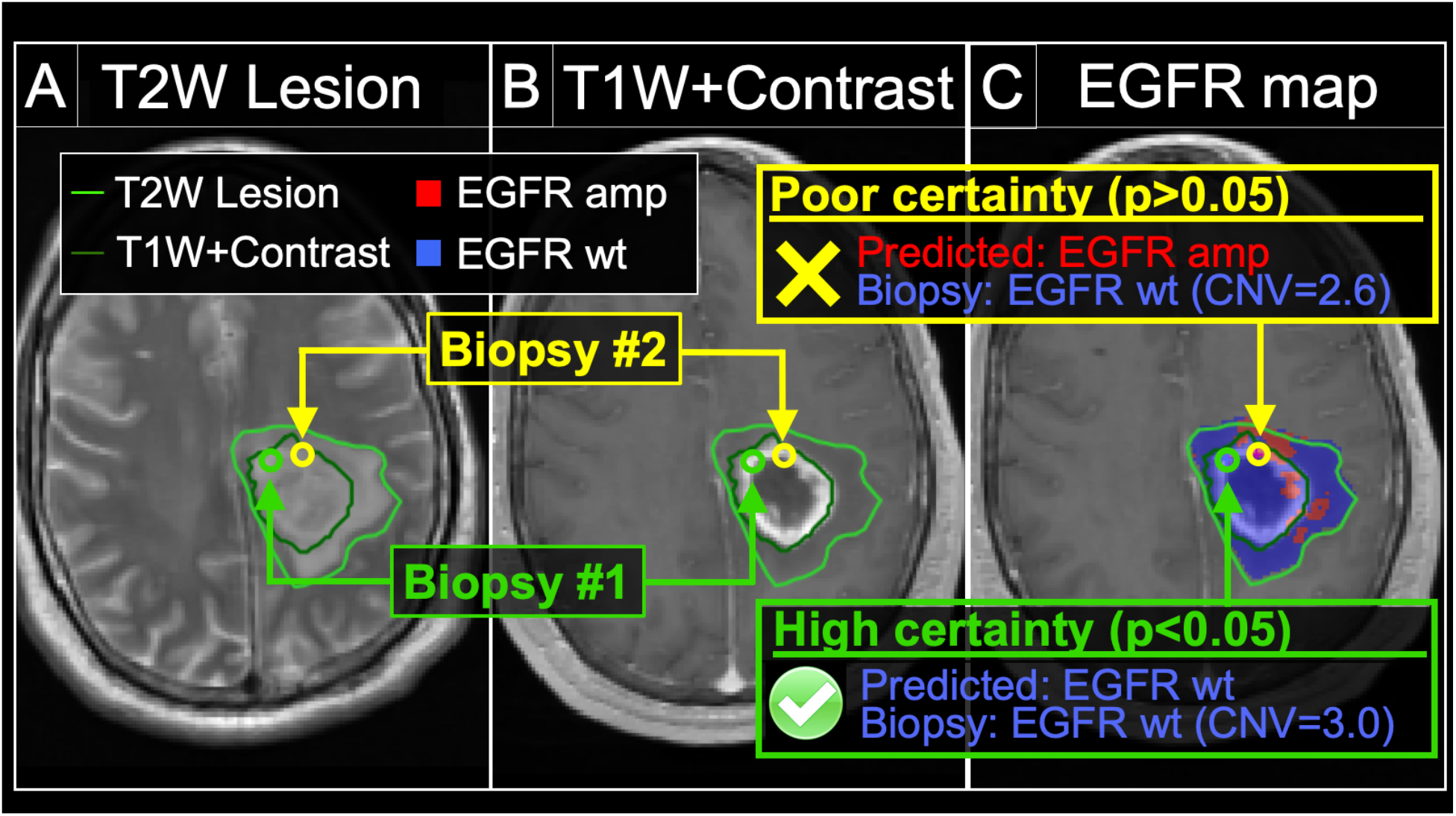
Model uncertainty can inform the likelihood of achieving an accurate radiogenomics prediction for EGFR amplification status. We obtained two separate biopsies (#1 and #2) from the same tumor in a 44 year-old male patient with primary GBM. The (A) T2W and (B) T1+C images demonstrate the enhancing (dark green outline, T1W+Contrast) and peripheral non-enhancing tumor segments (light green outline, T2W lesion). The (C) radiogenomics color map shows regions of predicted EGFR amplification (amp, red) and EGFR wildtype (wt, blue) status overlaid on the T1+C images. For biopsy #1 (green circle), the radiogenomics model predicted EGFR wildtype status (blue) with high certainty, which was concordant with biopsy results (green box). For biopsy #2 (yellow circle), the radiogenomics model showed poor certainty (i.e., high uncertainty), with resulting discordance between predicted (red) and actual EGFR status (yellow box).

## DISCUSSION

The era of genomic profiling has ushered new strategies for improving cancer care and patient outcomes through more personalized therapeutic approaches.^14^ In particular, the paradigm of individualized oncology can optimize targeted treatments to match the genetic aberrations and drug sensitivities for each tumor.^15^ This holds particular relevance for many aggressive tumors that may not be amenable to complete surgical resection, such as GBM or pancreatic adenocarcinoma, or other cancers that have widely metastasized. In these cases, the targeted drug therapies represent the primary lines of defense in combating the residual, unresected tumor populations that inevitably recur and lead to patient demise. Yet, a fundamental challenge for individualized oncology lies in our inability to resolve the internal genetic heterogeneity of these tumors. Taking GBM as an example, each patient tumor actually comprises multiple genetically distinct subpopulations, such that the genetic drug targets from one biopsy location may not accurately reflect those from other parts of the same tumor.^16^ Tissue sampling errors are magnified by the fact that surgical targeting favors MRI enhancing tumor components but leaves behind the residual subpopulations within the non-enhancing tumor segment.^17-19^ Ironically, while these uncharacterized residual subpopulations represent the primary targets of adjuvant therapy, their genetic drug targets remain “unknown”, even after gross total resection.^20,21^

With recent advances in the field of radiogenomics, image-based modeling offers a promising and clinically feasible solution to bridge the challenges of intratumoral heterogeneity. The past decade has seen a growing number of published studies comparing MRI with genetic profiles in GBM, typically in a non-localizing fashion.^21-29^ But for the first-time - based on work using machine-learning (ML) and image-localized biopsies - spatially resolved radiogenomics maps can now quantify the regional intratumoral variations of key GBM driver genes, at the image voxel level, throughout different regions of the same tumor.^18^ This includes highly relevant drug targets such as Epidermal Growth Factor Receptor (EGFR), which amplifies in as many as 30-45% of GBM tumors, and serves as the target for many clinically tested and commercially available inhibitors. Importantly, these radiogenomics maps of intratumoral heterogeneity offer a clinically viable solution for diagnosing the potentially unique drug targets within the residual unresected tumor segment, which otherwise represent a major barrier to individualized oncology.

As the data available to train machine learning models is never fully representative of the predictive cases, model predictions always have some inherent uncertainty in their predictions. To translate these models into clinical practice, the radiogenomics predictions need to not only be accurate, but must also inform the clinician of the confidence or certainty of each individual prediction. This underscores the importance of quantifying predictive uncertainty, but also highlights the major gaps that currently exist in this field. To date, no published studies have addressed predictive uncertainty in the context of radiogenomics. And yet, uncertainty represents a fundamental aspect of these data-driven approaches. In large part, this uncertainty stems from the inherent sparsity of training data, which limits the scope of the model domain. As such, there exists a certain degree of probability that any new unseen case may fall outside of the model domain, which would require the model to generalize or extrapolate beyond the domain to arrive at a prediction. We must recognize that all clinical models suffer from this sparsity of data, particularly when relying on spatially resolved biopsy specimens in the setting of intratumoral heterogeneity. But knowledge of which predicted cases fall within or outside of the model domain will help clinicians make informed decisions on whether to rely upon or disregard these model predictions on an individualized patient-by-patient basis.

The transductive learning GP model presented here addresses the challenges of predictive uncertainty at various levels. First, the model training incorporates the minimization of uncertainty as part of feature selection, which inherently optimizes the model domain to prioritize interpolation rather than extrapolation. Our data show that interpolated predictions generally correspond with higher predictive accuracy compared with extrapolated ones. Thus, by shifting the model domain towards interpolation, model performance rises. Second, along these same lines, the GP model can inform the likelihood of achieving an accurate prediction, by quantifying the level of uncertainty for each sample prediction. We observed a dramatic increase in classification accuracy among those sample predictions with low uncertainty (p<0.05), which corresponded with interpolated predictions. This allows the clinician to accept the reality that not all model predictions are correct. But by qualifying each sample prediction, the model can inform of which patient cases should be trusted over others. Third, the transductive learning process also appeared to impact the sample predictions suffering from border uncertainty, where the predicted distribution fell in close proximity to the classification threshold. Compared to the standard GP baseline model, the transductive learning model shifted the domain, such that most of these sample predictions could be interpolated, which further improved model performance. Finally, we noted that the incorporation of uncertainty in the training of the transductive learning model resulted in a substantial reduction in selected features and model complexity, compared with the standard GP approach to model training. While lower model complexity would suggest greater generalizability, further studies are likely needed to confirm this assertion.

We recognize various limitations to our study. First, our relatively small sample size precluded the separation of our cohort into training and validation sets. We employed leave-one-patient-out cross validation as a means to test our predictive models, but future studies are likely needed as we expand our patient cohort. We also acknowledge that there are various types of uncertainty that contribute to model performance that were not directly tested in this study. We focused on the most recognized source of uncertainty that impacts data-driven approaches like radiogenomics, which is data sparsity and the limitations of the model domain. But at the same time, we also designed patient recruitment, MRI scanning acquisition and post-processing methodology, surgical biopsy collection, and tissue treatment to minimize potential noise of data inputs. With that said, we understand that future work is needed to evaluate these potential sources of model uncertainty. Further, while neurosurgeons tried to minimize effects of brain shift by using small craniotomy sizes and by visually validating stereotactic biopsy locations, brain shift could have led to possible misregistration errors. Rigid-body coregistration of stereotactic imaging with the advanced MR-imaging was employed to also try to reduce possible geometric distortions. ^30,31^ However, our previous experience suggests that these potential contributions to misregistration results in about 1-2 mm of error, which is similar to previous studies using stereotactic needle biopsies.^32^ But as a potential source of uncertainty, future work can study these factors in a directed manner.

## CONCLUSION

In this study, we highlight the challenges of predictive uncertainty in radiogenomics and present a novel approach that not only quantifies model uncertainty, but also leverages it to enhance model performance and interpretability. This work offers a pathway to clinically integrating reliable radiogenomics predictions as part of decision support within the paradigm of individualized oncology.

## METHODS

### Acquisition and Processing of Clinical MRI and Histologic Data

#### Patient recruitment and Surgical biopsies

We recruited patients with clinically suspected GBM undergoing preoperative stereotactic MRI for surgical resection as previously described.^18^ we confirmed histologic diagnosis of GBM in all cases. Patients were recruited to this study, “Improving diagnostic Accuracy in Brain Patients Using Perfusion MRI,” under the protocol procedures approved by the Barrow Neurological Institute (BNI) institutional review board. Informed consent from each subject was obtained prior to enrollment. All data collection and protocol procedures were carried out following the approved guidelines and regulations outlined in the BNI IRB. Neurosurgeons used pre-operative conventional MRI, including T1-Weighted contrast-enhanced (T1+C) and T2-Weighted sequences (T2W), to guide multiple stereotactic biopsies as previously described.^18,33^ In short, each neurosurgeon collected an average of 3-4 tissue specimens from each tumor using stereotactic surgical localization, following the smallest possible diameter craniotomies to minimize brain shift. Neurosurgeons selected targets separated by at least 1 cm from both enhancing core (ENH) and non-enhancing T2/FLAIR abnormality in pseudorandom fashion, and recorded biopsy locations via screen capture to allow subsequent coregistration with multiparametric MRI datasets.

#### Histologic analysis and tissue treatment

Tissue specimens (target volume of 125mg) were flash frozen in liquid nitrogen within 1-2 min from collection in the operating suite and stored in −80°C freezer until subsequent processing. Tissue was retrieved from the freezer and embedded frozen in optimal cutting temperature (OCT) compound. Tissue was cut at 4 um sections in a −20 degree C cryostat (Microm-HM-550) utilizing microtome blade.^18,33^ Tissue sections were stained with hematoxylin and eosin (H&E) for neuropathology review to ensure adequate tumor content (≥50%).

### Whole Exome Sequencing and Alignment and Variant Calling

We performed DNA isolation and determined copy number variant (CNV) for all tissue samples using array comparative genomic hybridization (aCGH) and exome sequencing as previously published.^18,34-36^ This included application of previously described CNV detection to whole genome long insert sequencing data and exome sequencing.^18,34-36^ Briefly, tumor-normal whole exome sequencing was performed as previously described.^37^ DNA from fresh frozen tumor tissue and whole blood (for constitutional DNA analysis) samples were extracted and QC using DNAeasy Blood and Tissue Kit (Qiagen#69504). Tumor-normal whole exome sequencing was performed with the Agilent Library Prep Kit RNA libraries. Sequencing was performed using the Illumina HiSeq 4000 with 100bp paired-end reads. Fastq files were aligned with BWA 0.6.2 to GRCh37.62 and the SAM output were converted to a sorted BAM file using SAMtools 0.1.18. Lane level samples BAMs were then merged with Picard 1.65 if they were sequence across multiple lanes. Comparative variant calling for exome data was conducted with Seurat. For copy number detection was applied to the whole exome sequence as described (PMID 27932423). Briefly, copy number detection was based on a log2 comparison of normalized physical coverage (or clonal coverage) across tumor and normal whole exome sequencing data. Normal and tumor physical coverage was then normalized, smoothed and filtered for highly repetitive regions prior to calculating the log2 comparison. To quantify the copy number aberrations, CNV score was calculated based on the intensity of copy number change (log ratio).

#### MRI protocol, parametric maps, and image coregistration

**Conventional MRI and general acquisition conditions:** We performed all imaging at 3 Tesla field strength (Sigma HDx; GE-Healthcare Waukesha Milwaukee; Ingenia, Philips Healthcare, Best, Netherlands; Magnetome Skyra; Siemens Healthcare, Erlangen Germany) within 1 day prior to stereotactic surgery. Conventional MRI included standard pre- and post-contrast T1-Weighted (T1-C, T1+C, respectively) and pre-contrast T2-Weighted (T2W) sequences. T1W images were acquired using spoiled gradient recalled-echo inversion-recovery prepped (SPGR-IR prepped) (TI/TR/TE=300/6.8/2.8ms; matrix=320*224; FOV=26cm; thickness=2mm). T2W images were acquired using fast-spin-echo (FSE) (TR/TE=5133/78ms; matrix=320×192; FOV=26cm; thickness=2mm). T1+C images were acquired after completion of Dynamic Susceptibility-weighted Contrast-enhanced (DSC) Perfusion MRI (pMRI) following total Gd-DTPA (gadobenate dimeglumine) dosage of 0.15 mmol/kg as described below.^18,33,38^ **Diffusion Tensor (DTI)**: DTI imaging was performed using Spin-Echo Echo-planar imaging (EPI) [TR/TE 10000/85.2ms, matrix 256×256; FOV 30cm, 3mm slice, 30 directions, ASSET, B=0,1000]. The original DTI image DICOM files were converted to a FSL recognized NIfTI file format, using MRIConvert (http://lcni.uoregon.edu/downloads/mriconvert), before processing in FSL from semi-automated script. DTI parametric maps were calculated using FSL (http://fsl.fmrib.ox.ac.uk/fsl/fslwiki/), to generate whole-brain maps of mean diffusivity (MD) and fractional anisotrophy (FA) based on previously published methods.^20^ **DSC-pMRI**: Prior to DSC acquisition, preload dose (PLD) of 0.1 mmol/kg was administered to minimize T1W leakage errors. After PLD, we employed Gradient-echo (GE) EPI [TR/TE/flip angle=1500ms/20ms/6°, matrix 128×128, thickness 5mm] for 3 minutes. At 45 sec after the start of the DSC sequence, we administered another 0.05 mmol/kg i.v. bolus Gd-DTPA.^18,33,38^ The initial source volume of images from the GE-EPI scan contained negative contrast enhancement (i.e., susceptibility effects from the PLD administration) and provided the MRI contrast labeled EPI+C. At approximately 6 minutes after the time of contrast injection, the T2*W signal loss on EPI+C provides information about tissue cell density from contrast distribution within the extravascular, extracellular space.^33,39^ We performed leakage correction and calculated relative cerebral blood (rCBV) based on the entire DSC acquisition using IB Neuro (Imaging Biometrics, LLC) as referenced.^40,41^ We also normalized rCBV values to contralateral normal appearing white matter as previously described.^33,38^ **Image coregistration**: For image coregistration, we employed tools from ITK (www.itk.org) and IB Suite (Imaging Biometrics, LLC) as previously described.^18,33,38^ All datasets were coregistered to the relatively high quality DTI B0 anatomical image volume. This offered the additional advantage of minimizing potential distortion errors (from data resampling) that could preferentially impact the mathematically sensitive DTI metrics. Ultimately, the coregistered data exhibited in plane voxel resolution of ~1.17 mm (256×256 matrix) and slice thickness of 3mm.

#### ROI segmentation, Image feature extraction and Texture Analysis Pipeline

We generated regions of interest (ROIs) measuring 8×8×1 voxels (9.6×9.6×3mm) for each corresponding biopsy location. A board-certified neuroradiologist (L.S.H.) visually inspected all ROIs to ensure accuracy.^18,33^ From each ROI, we employed our in-house texture analysis pipeline to extract a total of 336 texture features from each sliding window. This pipeline, based on previous iterations^18,33^, included measurements of first-order statistics from raw image signals (18 features): mean (M) and standard deviation (SD) of gray-level intensities, Energy, Total Energy, Entropy, Minimum, 10th percentile, 90th percentile, Maximum, Median, Interquartile Range, Range, Mean Absolute Deviation (MAD), Robust Mean Absolute Deviation (rMAD), Root Mean Squared (RMS), Skewness, Kurtosis, Uniformity.^42^ We mapped intensity values within each window onto the range of 0–255. This step helped standardize intensities and reduced effects of intensity nonuniformity on features extracted during subsequent texture analysis. Texture analysis consisted of two separate but complementary texture algorithms: gray level co-occurrence matrix (GLCM),^43,44^ and Gabor Filters (GF)^45^, based on previous work showing high relevance to regional molecular and histologic characteristics.^18,33^ The output from the pipeline comprised a feature vector from each sliding window, composed of 56 features across each of the 6 MRI contrasts, for a total of 336 (6*56) features.

### Radiogenomics modeling and quantification of predictive uncertainty

#### Gaussian Process (GP) model (Standard)

A GP model was chosen to predict EGFR status using MRI features. A GP model offers the flexibility of identifying nonlinear relationships between input and output variables.^9,11–13^ More importantly, it generates a probability distribution for each prediction, which quantifies the uncertainty of the prediction. This capability is important for using the prediction result to guide clinical decision making. We applied this GP framework to our spatially matched MRI and image-localized genetic data to build a predictive model for the probability distributions of EGFR amplification for each biopsy specimen. Because the GP model generates probability distributions (rather than point estimates), this allows for direct quantification of both predictive uncertainty and predictive accuracy by using the predictive variance or standard deviation of the distribution and the predictive mean in comparison with the true EGFR status, respectively. The standard GP model measures but does not explicitly incorporate predictive uncertainties for model optimization during the training phase of development (as detailed below with Transductive Learning). In other words, the standard GP model can measure but is built without considering the predictive uncertainty.

Next we illustrate how GP works. Let {**x**_1_, *…*,**x***_N_*] be the input variables (i.e., texture features) corresponding to *N* samples. GP assumes a set of random functions corresponding to these samples, {*f*(**x**_1_)*,…,f*(**x***_N_*)}. This is called a Gaussian Process because any subset of {*f*(**x**_1_)*,…,f*(**x***_N_*)} follows a joint Gaussian distribution with zero mean and the covariance between two samples **x***_i_* and **x***_j_* computed by a kernel function *K*(**x***_i_*, **x***_j_*). The input variables are linked with the output (i.e., the transformed CNV) by *y_i_ = f*(**x***_i_*) *+ ∊_i_*, where *∊_i_ ~N*(0*,σ*^2^) is a Gaussian noise.

Given a training dataset {**X,Y**} where **X** is a matrix containing the input variables of all training samples and **Y** is a vector containing the output variables of these samples, the predictive distribution for a new test sample **x***\** is

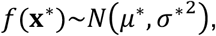

where

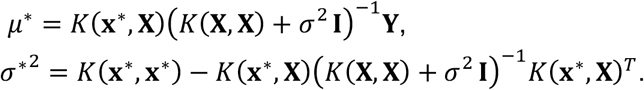

##### Uncertainty quantification

While the predictive mean can be used as a point estimate of the prediction, the variance provides additional information about the uncertainty of the prediction. Furthermore, using this predictive distribution, one can test the hypothesis that a prediction is greater or less than a cutoff of interest. For example, in our case, we are interested in knowing if the prediction of the CNV of a sample is greater than 3.5 (considered as amplification). A small p value (e.g., <0.05) of the hypothesis testing implies prediction with certainty.

##### Feature selection

When the input variables are high-dimensional, including all of them in training a GP model has the risk of overfitting. Therefore, feature selection is needed. We used forward stepwise selection^46^, which started with an empty feature set and added one feature at each step that maximally improves a pre-defined criterion until no more improvement can be achieved. To avoid overfitting, a commonly used criterion is the accuracy computed on a validation set; when the sample size is limited, cross-validation accuracy can be adopted. In our case, we adopt leave-one-patient-out cross validation accuracy to be consistent with the natural grouping of samples. The accuracy is computed by comparing the true and predicted CNVs; a match is counted when both are on the same side of 3.5 (i.e., both being amplified or non-amplified).

### Transductive Learning GP model

We sought to develop a radiogenomics model that would not only maximize predictive accuracy but also minimize the predictive uncertainty. To this end, we added a Transductive Learning component to the standard GP model (described above), which quantifies predictive uncertainty, and uses that information during the model training process to minimize uncertainty on subsequent predictions. Briefly, this model employs Transductive Learning to perform feature selection in an iterative, stepwise fashion to prioritize the minimization of average predictive uncertainty during model training. We applied this GP model with Transductive Learning to our spatially matched MRI and image-localized genetic data to predict the probability distribution of EGFR amplification for each biopsy specimen.

Although a typical supervised learning model is trained using labeled samples, i.e., samples with both input and output variables available, some machine learning algorithms have been developed to additionally incorporate unlabeled samples, i.e., samples with only input variables available. These algorithms belong to a machine learning subfield called transductive learning or semi-supervised learning.^47^ Transductive learning is beneficial when sample labeling is costly or labor-intensive, which results in a limited sample size. This is the case for our problem.

While the standard GP model described in the previous section can only utilize labeled samples, a transductive GP model was developed by Wang et al.^48^ Specifically, let {**X***_L_*, **Y***_L_*) and {**X***_U_*} be the sets of labeled and unlabeled samples used in training, respectively. A transductive GP first generates predictions for the unlabeled samples by applying the standard GP to the labeled samples, 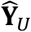. Here 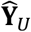 contains the means of the predictive distributions. Then, a combined dataset 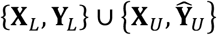 was used as the training dataset to generate a predictive distribution for each new test sample **x**^*^, i.e.,

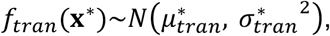

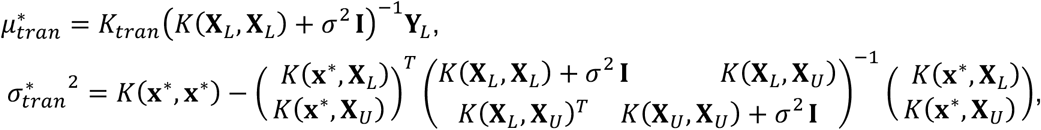

where

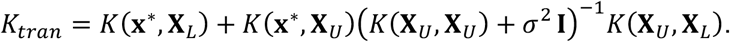

To decide which unlabeled samples to include in transductive GP, Wang et al.^48^ points out the importance of “self-similarity”, meaning the similarity between the unlabeled and test samples. Based on this consideration, we included the eight neighbors of the test sample on the image as the unlabeled samples. The labeled samples are those from other patients (i.e., other than the patient where the test sample is from), which are the same as the samples used in GP.

#### Uncertainty quantification

Because transductive GP also generates a predictive distribution, the same approach as that described for GP can be used to quantify the uncertainty of the prediction.

#### Feature selection

The same forward stepwise selection procedure as GP was adopted to select features for transductive GP. Because transductive GP significantly reduces the prediction variance, we found that feature selection could benefit from minimizing leave-one-patient-out cross validation uncertainty as the criterion instead of maximizing the accuracy. Specifically, we computed the p value for each prediction that reflects the uncertainty (the smaller the p, the more certain of the prediction) and selected features to minimize the leave-one-patient-out cross validation p value.

### Model comparison by theoretical analysis

The original paper that proposed transductive GP showed empirical evidence that it outperformed GP but not theoretical justification. In this paper, we derived several theorems to reveal the underlying reasons. The proofs are skipped but available upon request.

#### Theorem 1

When applying both GP and transdudtive GP to predict a test sample **x**^*^, the predictive variance of transductive GP is no greater than GP, i.e., 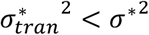.

#### Theorem 2

Consider a test sample **x***^*^*. Let {**X***_L_*, **Y***_L_*} be the set of labeled samples used in training by GP. Let {**X***_U_*} be the set of unlabeled samples used in training by transductive GP in addition to the labeled set. If *K*(**X***_U_*,**X***_L_*) *→* **0** and (**X***_U_*,**x***^*^*) *→* **0**, i.e., the distances of the unlabeled samples with respect to the labeled and test samples go to zero, then the predictive distribution for **x***^*^* by transductive GP, *f_tran_*(**x**^*^), converges to that by GP, *f*(**x**^*^), with respect to Kullback–Leibler divergence, i.e., 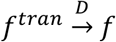.

### Model comparison on prediction accuracy and uncertainty

Using the selected features from each model (GP and transductive GP), we computed the prediction accuracy and uncertainty under leave-one-patient-out cross validation (LopoCV). The output from each GP model comprised a predictive distribution including a predictive mean and a predictive variance. We used the predictive mean as the point estimator for the CNV on the transformed scale, and used this to classify each biopsy sample as either EGFR amplified (CNV>3.5) or EGFR non-amplified (CNV≤3.5). This process was iterated until every patient served as the test case (all other patients as training). Note that LopoCV in theory provides greater rigor compared to k-fold cross validation or leave-one-out cross validation (LOOCV), which leaves out a single biopsy sample as the test case. LopoCV would likely better simulate clinical practice (i.e., the model is used on a per-patient basis, rather than on a per-sample basis). In addition to predictive mean, each GP model output also includes predictive variance for each sample, which allows for quantification of predictive uncertainty. Specifically, for each prediction on each biopsy, we tested the hypothesis that the sample belongs to the class predicted by the mean (H1) versus not (H0), using a standard one-sided z test. The p-value of this test reflects the certainty of the prediction, such that smaller p values correspond with lower predictive uncertainty (i.e., greater certainty) for each sample classified by the model.

#### Classification of EGFR status and CNV data transformation

We employed the CNV threshold of 3.5 to classify each biopsy sample as EGFR normal (CNV≤3.5) vs EGFR amplified (CNV>3.5) as a statistically conservative approach to differentiating amplified samples from diploidy and triploidy samples.^49^ As shown in prior work, the log-scale CNV data for EGFR status can also exhibit heavily skewed distributions across a population of biopsy samples, which can manifest as a long tail with extremely large values (up to 22-fold log scale increase) in a relative minority of EGFR amplified samples.^50^ Such skewed distributions can present challenges for ML training, which we addressed using data transformation.^51,52^ This transformation maintained identical biopsy sample ordering between transformed and original scales, but condensed the spacing between samples with extreme values on the transformed scale, such that the distribution width of samples with CNV>3.5 approximated that of the samples with CNV≤3.5.

#### Leave-one-patient-out-Cross-Validation (LopoCV) and quantification of Predictive Uncertainty

To determine predictive accuracies for each GP model (without vs with Transductive Learning), we employed LopoCV. In this scheme, one randomly selected patient (and all of their respective biopsy samples) served as the test case, while the other remaining patients (and their biopsy data) served to train the model. Training consisted of fitting a GP regression to the entire training data set, and then using the trained GP regression model to predict all of the samples from the test patient case. The output from each GP model comprised a predictive distribution on each biopsy sample of the test patient, including a predictive mean and a predictive variance. We used the predictive mean as the point estimator for the CNV on the transformed scale, and used this to classify each biopsy sample as either EGFR amplified (CNV>3.5) or EGFR non-amplified (CNV≤3.5). This process was iterated until every patient served as the test case. Note that LopoCV in theory provides greater rigor compared to k-fold cross validation or leave-one-out cross validation (LOOCV), which leaves out a single biopsy sample as the test case. LopoCV would likely better simulate clinical practice (i.e., the model is used on a per-patient basis, rather than on a per-sample basis). In addition to predictive mean, each GP model output also includes predictive variance for each sample, which allows for quantification of predictive uncertainty. Specifically, for each prediction on each biopsy, we tested the hypothesis that the sample belongs to the class predicted by the mean (H1) versus not (H0), using a standard one-sided z test. The p-value of this test reflects the certainty of the prediction, such that smaller p values correspond with lower predictive uncertainty (i.e., greater certainty) for each sample classified by the model. We prioritized the lowest predictive uncertainty as those predictions with the lowest range of p-values (p<0.05). We also evaluated incremental ranges of p-values (e.g., <0.10; <0.15, etc) as gradations of progressively decreasing predictive uncertainty.

#### Subject Population, Clinical Data, and Genomic Analysis

We recruited a total of 25 untreated GBM patients and collected 95 image-localized biopsy samples for analysis. Patient demographics and clinical data are summarized in **Table 3**. The number of biopsies ranged from 1 to 6 per patient. Measured log-scale CNV status of EGFR amplification ranged from 1.876 to 82.43 across all 95 biopsy samples.

**Table 3:** Patient demographics for the 25 untreated glioblastoma (GBM) tumors. Patients labeled A through Y. M=male, F=female. Patient L on histology was diagnosed as Grade III Anaplastic Astrocytoma, although molecular features were consistent with Glioblastoma (e.g., EGFR amplification). NA = not available. Wt = wild type. aCGH = array comparative genomic hybridization; WES = whole exome sequencing

| Patient | Sex | Age (yrs) | Type | Pathology | IDH.status | Sequenced |
| --- | --- | --- | --- | --- | --- | --- |
| A | M | 62 | glioblastoma | primary | NA | aCGH |
| B | M | 66 | glioblastoma | primary | NA | aCGH |
| C | M | 23 | glioblastoma | primary | NA | aCGH |
| D | M | 47 | glioblastoma | primary | NA | aCGH |
| E | F | 78 | glioblastoma | primary | NA | aCGH |
| F | M | 80 | glioblastoma | primary | NA | aCGH |
| G | F | 78 | glioblastoma | primary | NA | aCGH |
| H | M | 71 | glioblastoma | primary | NA | aCGH |
| I | M | 77 | glioblastoma | primary | NA | aCGH |
| J | M | 31 | glioblastoma | primary | NA | aCGH |
| K | F | 85 | glioblastoma | primary | NA | aCGH |
| L | M | 65 | Anaplastic Astrocytoma | primary | NA | aCGH |
| M | F | 40 | glioblastoma | primary | NA | aCGH |
| N | F | 51 | glioblastoma | primary | wt | WES |
| O | M | 76 | glioblastoma | primary | wt | WES |
| P | M | 61 | glioblastoma | primary | wt | WES |
| Q | M | 67 | glioblastoma | primary | V178I/G105G | WES |
| R | M | 44 | glioblastoma | primary | wt | WES |
| S | F | 53 | glioblastoma | primary | R132H | WES |
| T | F | 62 | glioblastoma | primary | wt | WES |
| U | F | 61 | glioblastoma | primary | NA | aCGH |
| V | F | 78 | glioblastoma | primary | wt | WES |
| W | F | 75 | glioblastoma | primary | wt | WES |
| X | F | 52 | glioblastoma | primary | V178I | WES |
| Y | F | 69 | glioblastoma | primary | wt | WES |

## Data Availability

Data will be made available upon publication in a peer reviewed journal.

